# Protection of SARS-CoV-2 natural infection against reinfection with the Omicron BA.4 or BA.5 subvariants

**DOI:** 10.1101/2022.07.11.22277448

**Authors:** Heba N. Altarawneh, Hiam Chemaitelly, Houssein H. Ayoub, Mohammad R. Hasan, Peter Coyle, Hadi M. Yassine, Hebah A. Al-Khatib, Fatiha M. Benslimane, Zaina Al-Kanaani, Einas Al-Kuwari, Andrew Jeremijenko, Anvar Hassan Kaleeckal, Ali Nizar Latif, Riyazuddin Mohammad Shaik, Hanan F. Abdul-Rahim, Gheyath K. Nasrallah, Mohamed Ghaith Al-Kuwari, Adeel A. Butt, Hamad Eid Al-Romaihi, Mohamed H. Al-Thani, Abdullatif Al-Khal, Roberto Bertollini, Patrick Tang, Laith J. Abu-Raddad

## Abstract

This study estimates the effectiveness of previous infection with SARS-CoV-2 in preventing reinfection with Omicron BA.4/BA.5 subvariants using a test-negative, case–control study design. Cases (SARS-CoV-2-positive test results) and controls (SARS-CoV-2-negative test results) were matched according to sex, 10-year age group, nationality, comorbid condition count, calendar week of testing, method of testing, and reason for testing. Effectiveness was estimated using the S-gene “target failure” (SGTF) infections between May 7, 2022-July 4, 2022. SGTF status provides a proxy for BA.4/BA.5 infections, considering the negligible incidence of other SGTF variants during the study. Effectiveness was also estimated using all diagnosed infections between June 8, 2022-July 4, 2022, when BA.4/BA.5 dominated incidence. Effectiveness of a previous pre-Omicron infection against symptomatic BA.4/BA.5 reinfection was 15.1% (95% CI: -47.1-50.9%), and against any BA.4/BA.5 reinfection irrespective of symptoms was 28.3% (95% CI: 11.4-41.9%). Effectiveness of a previous Omicron infection against symptomatic BA.4/BA.5 reinfection was 76.1% (95% CI: 54.9-87.3%), and against any BA.4/BA.5 reinfection was 79.7% (95% CI: 74.3-83.9%). Results using all diagnosed infections when BA.4/BA.5 dominated incidence confirmed the same findings. Sensitivity analyses adjusting for vaccination status confirmed study results. Protection of a previous infection against BA.4/BA.5 reinfection was modest when the previous infection involved a pre-Omicron variant, but strong when the previous infection involved the Omicron BA.1 or BA.2 subvariants. Protection of a previous infection against BA.4/BA.5 was lower than that against BA.1/BA.2, consistent with BA.4/BA.5’s greater capacity for immune-system evasion than that of BA.1/BA.2.

## Introduction

Omicron BA.4 and BA.5 (B.1.1.529) subvariants of the severe acute respiratory syndrome coronavirus 2 (SARS-CoV-2) have emerged in recent weeks and exhibit substantial capacity to escape from neutralizing antibodies.^1^ These subvariants were introduced in Qatar by early May of 2022 (Figure 1), and became the dominant subvariants by June 8, 2022 (Figure 2). We estimated effectiveness of previous infection in preventing reinfection with BA.4/BA.5 using a test-negative, case–control study design.^2^

**Figure 1.**
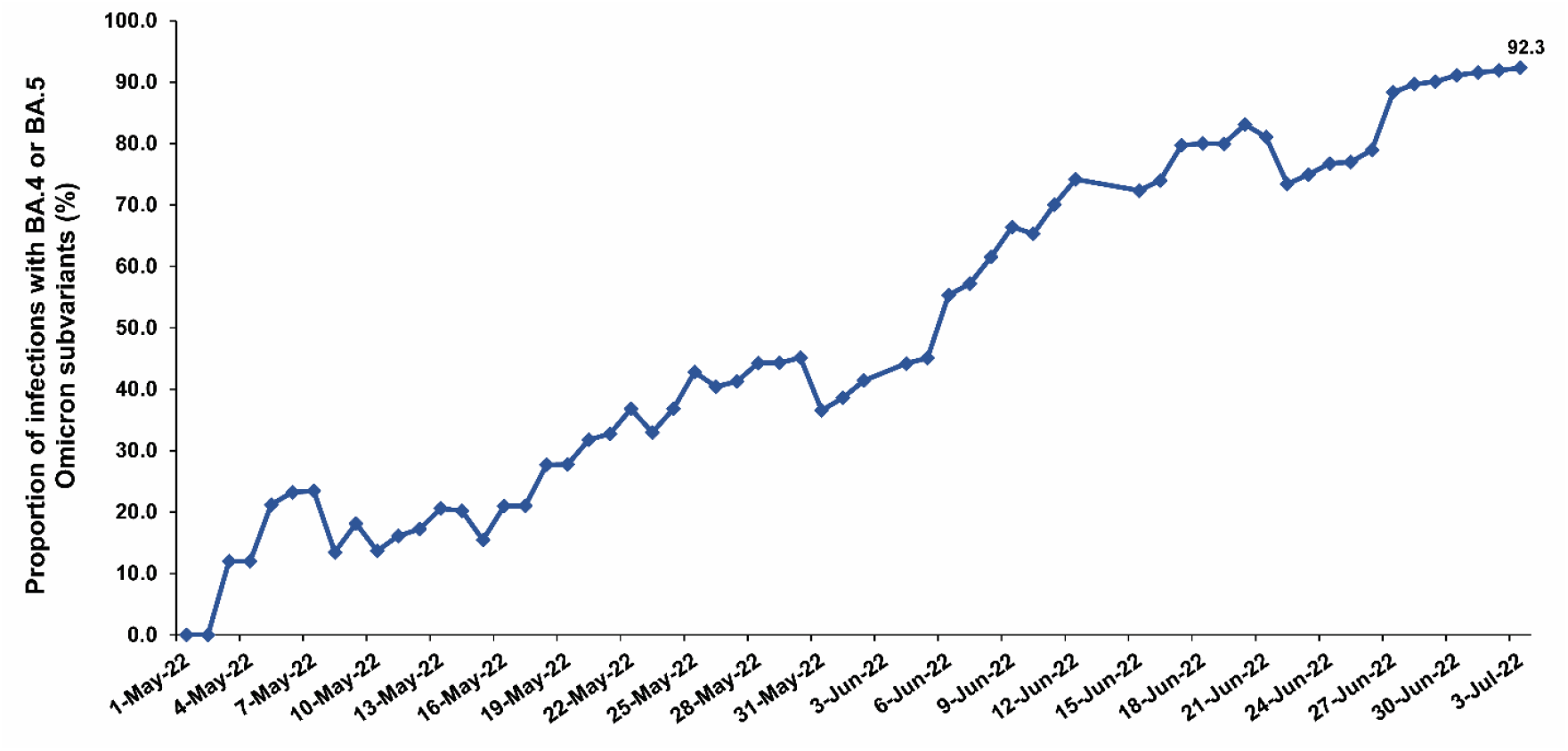
Proportion of SARS-CoV-2 Omicron infections that are with the BA.4 or BA.5 subvariants versus with the BA.2 subvariant, the only other subvariant of appreciable presence in Qatar between May 1, 2022 and July 3, 2022. BA.4 or BA.5 subvariant status was proxied as an S-gene “target failure” (SGTF) status in the PCR testing conducted using the TaqPath COVID-19 Combo Kit (Thermo Fisher Scientific, USA^24^).

**Figure 2.**
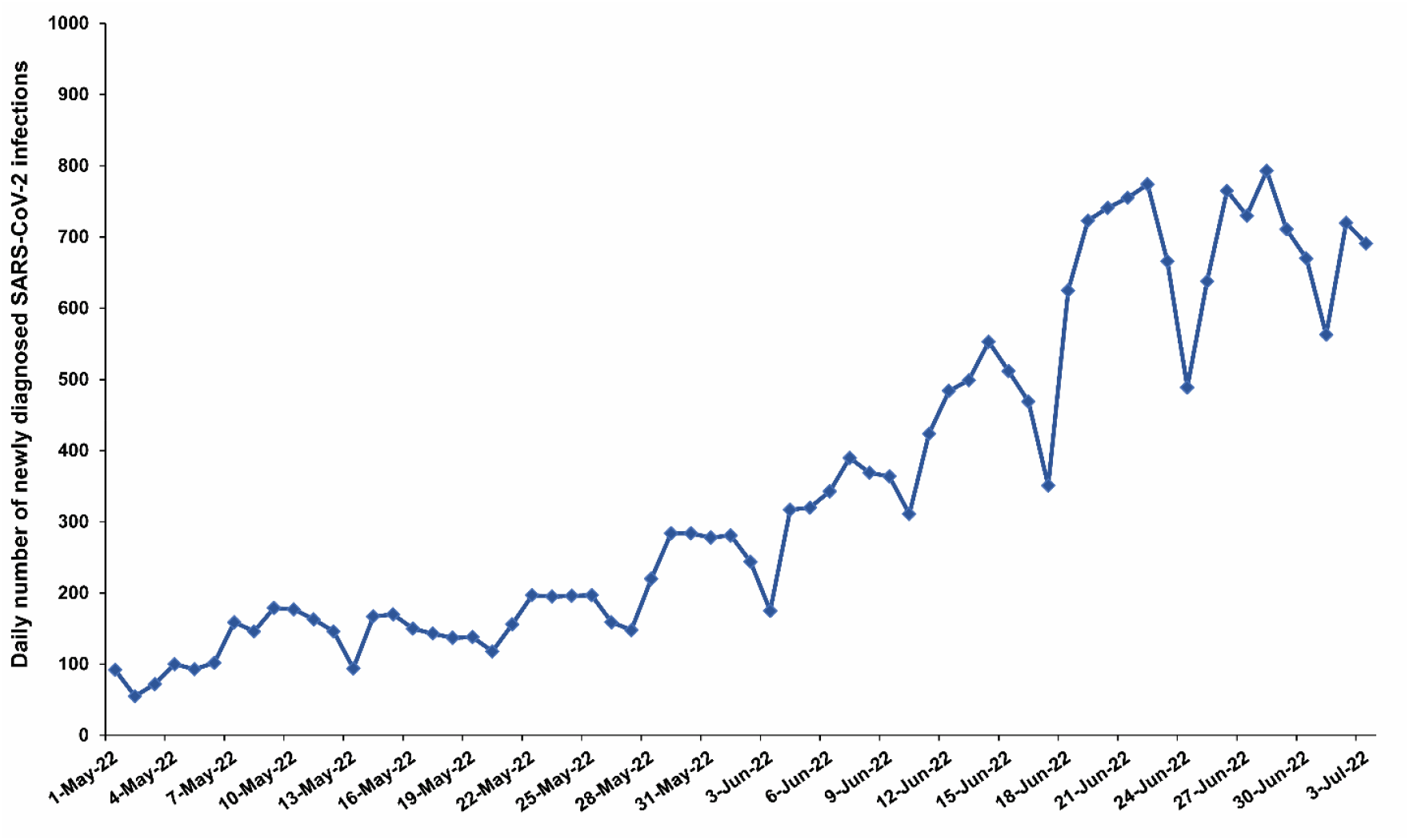
Daily number of newly diagnosed SARS-CoV-2 infections between May 1, 2022 and July 3, 2022.

## Methods

### Study population, data sources, and study design

This study was conducted in the resident population of Qatar, applying the test-negative, case-control study design^2-4^ to investigate the protection afforded by previous SARS-CoV-2 infection in preventing reinfection with SARS-CoV-2 BA.4 or BA.5 Omicron^5^ subvariants. Effectiveness of previous infection in preventing reinfection (*PE*_*S*_) was defined as the proportional reduction in susceptibility to infection among those with previous infection versus those without.^2,6^ The test-negative methodology was recently developed and validated for the specific derivation of rigorous and robust estimates for SARS-CoV-2 *PE*_*S*_,^2^ and has been applied in other recent studies for this purpose.^7-9^

The study analyzed the national, federated databases for coronavirus disease 2019 (COVID-19) laboratory testing, vaccination, clinical infection data, hospitalization, and death, retrieved from the integrated nationwide digital-health information platform. Databases include all SARS-CoV-2-related data and associated demographic information, with no missing information, since pandemic onset, documenting all polymerase chain reaction (PCR) testing and more recently, rapid antigen (RA) testing conducted at healthcare facilities (from January 5, 2022 onward). Every PCR test (but not every RA test) conducted in Qatar is classified on the basis of symptoms and the reason for testing (clinical symptoms, contact tracing, surveys or random testing campaigns, individual requests, routine healthcare testing, pre-travel, at port of entry, or other). PCR and RA testing in Qatar is done at a mass scale, where about 5% of the population are tested every week.^10^ Most infections are diagnosed not because of appearance of symptoms, but because of routine testing.^10^ Qatar has unusually young, diverse demographics, in that only 9% of its residents are ≥50 years of age, and 89% are expatriates from over 150 countries.^11,12^ Qatar launched its COVID-19 vaccination program in December of 2020 using BNT162b2 and mRNA-1273 vaccines.^13^ Further descriptions of the study population and these national databases have been reported previously.^10,12-17^

For estimation of *PE*_*S*_ against BA.4 or BA.5 infection, we exact-matched cases (SARS-CoV-2-positive tests) and controls (SARS-CoV-2-negative tests) in a one-to-five ratio by sex, 10-year age group, nationality, comorbid condition count, calendar week of testing, method of testing (PCR or RA), and reason for testing. Matching was done to control for known differences in risk of exposure to SARS-CoV-2 infection in Qatar.^12,18-21^ Matching by these factors was previously shown to provide adequate control of differences in risk of SARS-CoV-2 exposure in studies of different designs and that included control groups to test for null effects, including test-negative case-control studies.^10,13,16,22,23^

Effectiveness was estimated using two sets of cases. The first one includes the S-gene “target failure” (SGTF) cases identified using the TaqPath COVID-19 Combo Kit (Thermo Fisher Scientific, USA^24^). Routine PCR mass testing was designed to allow capture of SGTF variants during the study. SGTF status provides a proxy for BA.4 or BA.5 cases, considering the limited incidence of other variants with SGTF in Qatar during the study, between May 7, 2022 and July 4, 2022. To improve on statistical precision, an additional estimate was generated using all cases diagnosed when BA.4 and BA.5 dominated incidence, that is between June 8, 2022 and July 4, 2022. This estimate provides an upper bound for the effectiveness considering that some infections during this time could have been with BA.2, a subvariant associated with less immune evasion compared to BA.4 and BA.5.^1,8^

Only the first SARS-CoV-2-positive test for an individual occurring during the study period was included, while all SARS-CoV-2-negative tests were included. Controls included SARS-CoV-2-negative tests on individuals with no record of a SARS-CoV-2-positive test during the study.

SARS-CoV-2 reinfection is conventionally defined as a documented infection ≥90 days after an earlier infection, to avoid misclassification of prolonged PCR positivity as reinfection, if a shorter time interval is used.^8,25^ Previous infection was thus defined as a SARS-CoV-2-positive test ≥90 days before this study’s SARS-CoV-2 test. Cases or controls with SARS-CoV-2-positive tests <90 days before the study’s SARS-CoV-2 test were excluded.

Every control that met the inclusion criteria and that could be matched to a case was included in analysis. The above inclusion and exclusion criteria were implemented to minimize different types of potential bias, as informed by previous analyses.^7,8,10^ Previous infections were classified as pre-Omicron versus Omicron previous infections based on whether they occurred before or after the Omicron wave that started in Qatar on December 19, 2021.^7,8,17^

## Laboratory methods

### Real-time reverse-transcription polymerase chain reaction testing

Nasopharyngeal and/or oropharyngeal swabs were collected for PCR testing and placed in Universal Transport Medium (UTM). Aliquots of UTM were: 1) extracted on KingFisher Flex (Thermo Fisher Scientific, USA), MGISP-960 (MGI, China), or ExiPrep 96 Lite (Bioneer, South Korea) followed by testing with real-time reverse-transcription PCR (RT-qPCR) using TaqPath COVID-19 Combo Kits (Thermo Fisher Scientific, USA) on an ABI 7500 FAST (Thermo Fisher Scientific, USA); 2) tested directly on the Cepheid GeneXpert system using the Xpert Xpress SARS-CoV-2 (Cepheid, USA); or 3) loaded directly into a Roche cobas 6800 system and assayed with the cobas SARS-CoV-2 Test (Roche, Switzerland). The first assay targets the viral S, N, and ORF1ab gene regions. The second targets the viral N and E-gene regions, and the third targets the ORF1ab and E-gene regions.

All PCR testing was conducted at the Hamad Medical Corporation Central Laboratory or Sidra Medicine Laboratory, following standardized protocols.

### Rapid antigen testing

SARS-CoV-2 antigen tests were performed on nasopharyngeal swabs using one of the following lateral flow antigen tests: Panbio COVID-19 Ag Rapid Test Device (Abbott, USA); SARS-CoV-2 Rapid Antigen Test (Roche, Switzerland); Standard Q COVID-19 Antigen Test (SD Biosensor, Korea); or CareStart COVID-19 Antigen Test (Access Bio, USA). All antigen tests were performed point-of-care according to each manufacturer’s instructions at public or private hospitals and clinics throughout Qatar with prior authorization and training by the Ministry of Public Health (MOPH). Antigen test results were electronically reported to the MOPH in real time using the Antigen Test Management System which is integrated with the national COVID-19 database.

### Viral genome sequencing and classification of infections by variant type

Surveillance for SARS-CoV-2 variants in Qatar is based on viral genome sequencing and multiplex RT-qPCR variant screening^26^ of random positive clinical samples,^10,15,16,27-29^ complemented by deep sequencing of wastewater samples.^27,30^ Further details on the viral genome sequencing and multiplex RT-qPCR variant screening throughout the SARS-CoV-2 waves in Qatar can be found in previous publications.^7,8,10,15-17,27-29,31-33^

A total of 82 random SARS-CoV-2-positive specimens collected between May 28, 2022 and June 10, 2022 were viral whole-genome sequenced on a Nanopore GridION sequencing device. Of these, 1 (1.2%) was confirmed as Omicron BA.1, 38 (46.3%) as BA.2, 9 (11.0%) as BA.4, and 34 (41.5%) as BA.5.

Moreover, between June 5, 2022 and June 25, 2022, whole-genome sequencing of an additional 93 random SARS-CoV-2-positive specimens with PCR cycle threshold (Ct) values ≤25 showed 63 (67.7%) were BA.5, 7 (7.5%) were BA.4, 19 (20.4%) were BA.2, and 4 (4.3%) were BA.1. Additionally, between May 1, 2022 and June 15, 2022, S-gene sequencing of 84 other random SARS-CoV-2-positive specimens with SGTF showed 81 (96.4%) were BA.4/BA.5 and 3 (3.6%) were BA.1.

### Statistical analysis

While all records of SARS-CoV-2 testing were examined for selection of cases and controls, only matched samples were analyzed. Cases and controls were described using frequency distributions and measures of central tendency and compared using standardized mean differences. A standardized mean difference was defined as the difference in the mean of a covariate between groups, divided by the pooled standard deviation, with values <0.1 indicating optimal matching.^34^

*PE*_*S*_ was derived as one minus the ratio of the odds of previous infection in cases (SARS-CoV-2-positive tests), to the odds of previous infection in controls (SARS-CoV-2-negative tests):^2^

*PE*_*S*_ = 1 − odds ratio of prior infection among cases versus controls.

Odds ratios and associated 95% confidence intervals (CIs) were derived using conditional logistic regression, factoring the matching in the study design. This analytical approach, that also factors matching by calendar week of test, minimizes potential bias due to variation in epidemic phase^3,35^ and roll-out of vaccination during the study.^3,35^ CIs were not adjusted for multiplicity and thus should not be used to infer definitive differences between different groups. Interactions were not investigated.

Sensitivity analyses were conducted to assess the robustness of estimates of *PE*_*S*_. This was done by additionally adjusting for vaccination status in the conditional logistic regression. Statistical analyses were conducted in STATA/SE version 17.0 (Stata Corporation, College Station, TX, USA).

### Oversight

Hamad Medical Corporation and Weill Cornell Medicine-Qatar Institutional Review Boards approved this retrospective study with a waiver of informed consent. The study was reported following the Strengthening the Reporting of Observational Studies in Epidemiology (STROBE) guidelines. The STROBE checklist is found in Table 1.

**Table 1.**
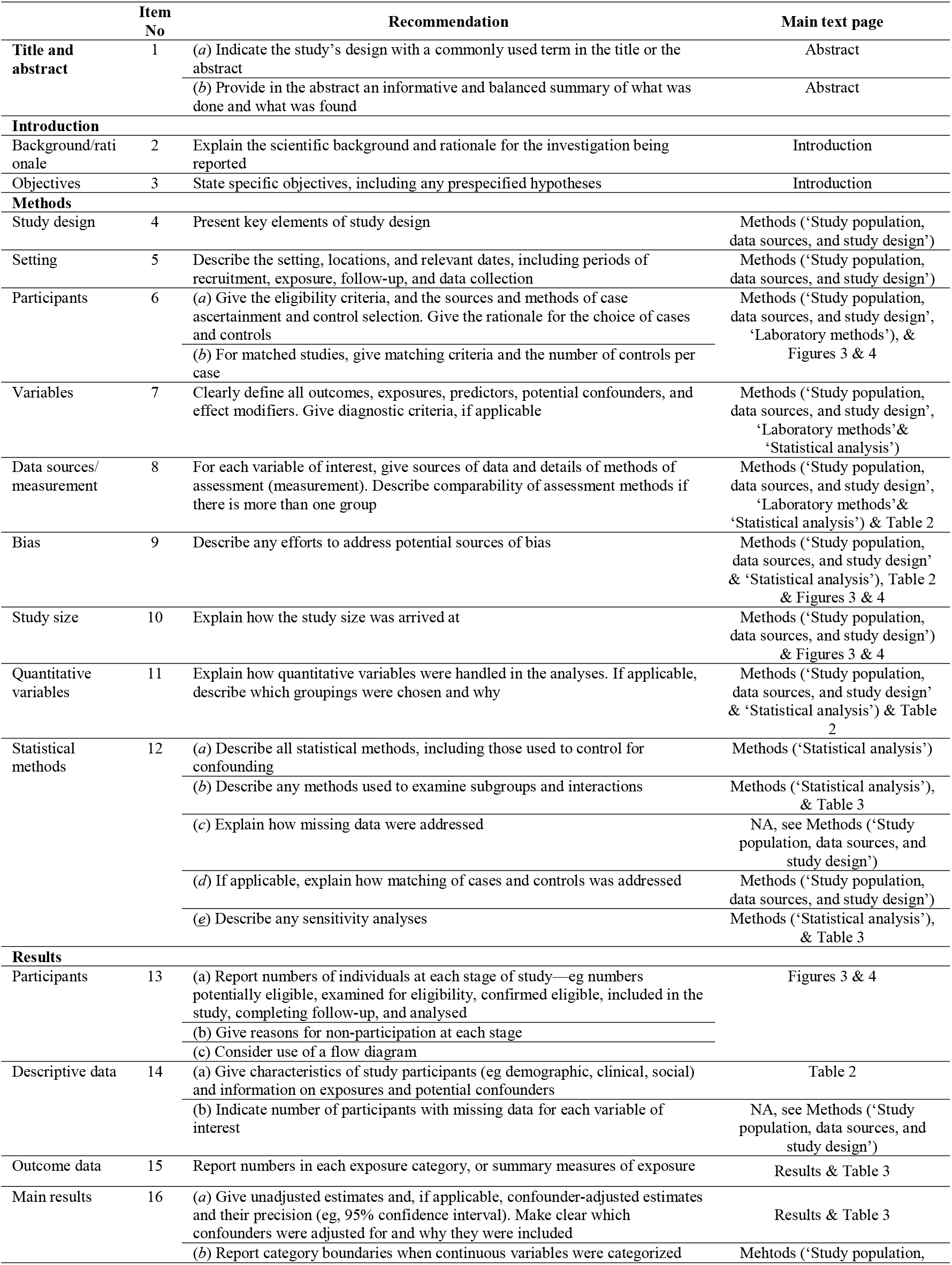
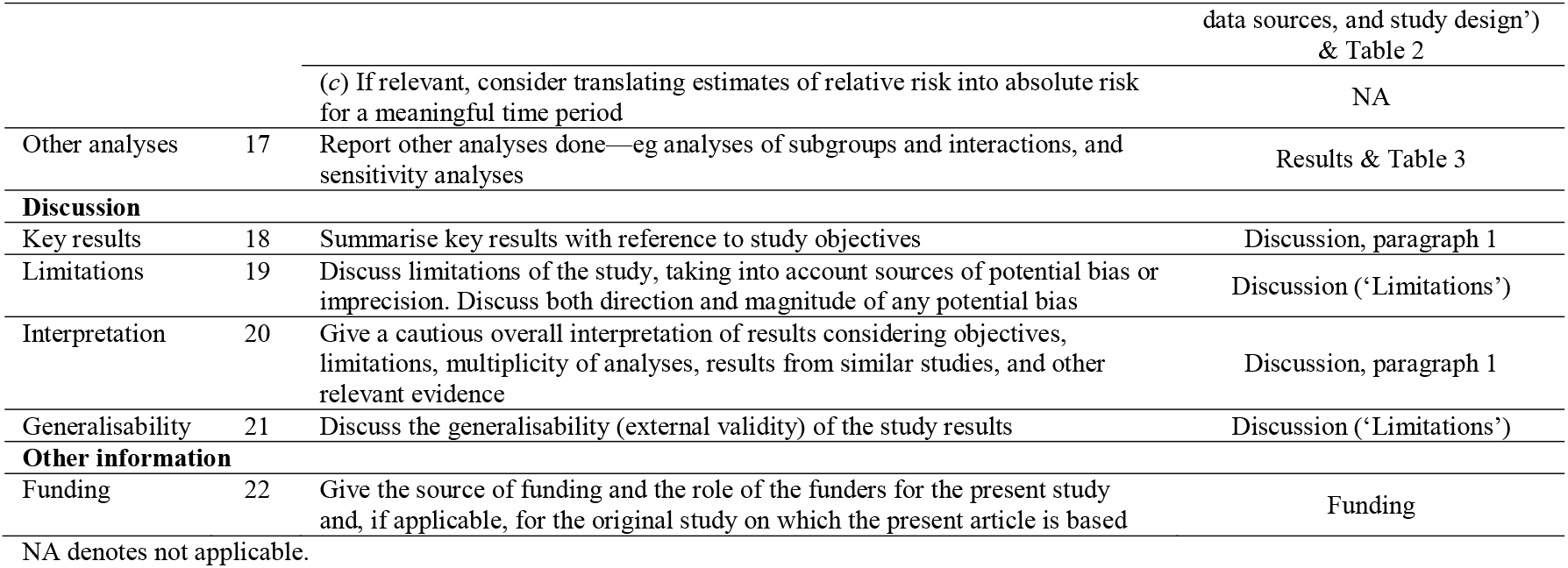
STROBE checklist for case-control studies.

|  | Item No | Recommendation | Main text page |
| --- | --- | --- | --- |
| Title and abstract | 1 | (a) Indicate the study’s design with a commonly used term in the title or the abstract | Abstract |
|  |  | (b) Provide in the abstract an informative and balanced summary of what was done and what was found | Abstract |
| Introduction |  |  |  |
| Background/rationale | 2 | Explain the scientific background and rationale for the investigation being reported | Introduction |
| Objectives | 3 | State specific objectives, including any prespecified hypotheses | Introduction |
| Methods |  |  |  |
| Study design | 4 | Present key elements of study design | Methods (‘Study population, data sources, and study design’) |
| Setting | 5 | Describe the setting, locations, and relevant dates, including periods of recruitment, exposure, follow-up, and data collection | Methods (‘Study population, data sources, and study design’) |
| Participants | 6 | (a) Give the eligibility criteria, and the sources and methods of case ascertainment and control selection. Give the rationale for the choice of cases and controls<br>(b) For matched studies, give matching criteria and the number of controls per case | Methods (‘Study population, data sources, and study design’, ‘Laboratory methods’), & Figures 3 & 4 |
| Variables | 7 | Clearly define all outcomes, exposures, predictors, potential confounders, and effect modifiers. Give diagnostic criteria, if applicable | Methods (‘Study population, data sources, and study design’, ‘Laboratory methods’ & ‘Statistical analysis’) |
| Data sources/measurement | 8 | For each variable of interest, give sources of data and details of methods of assessment (measurement). Describe comparability of assessment methods if there is more than one group | Methods (‘Study population, data sources, and study design’, ‘Laboratory methods’ & ‘Statistical analysis’) & Table 2 |
| Bias | 9 | Describe any efforts to address potential sources of bias | Methods (‘Study population, data sources, and study design’ & ‘Statistical analysis’), Table 2 & Figures 3 & 4 |
| Study size | 10 | Explain how the study size was arrived at | Methods (‘Study population, data sources, and study design’) & Figures 3 & 4 |
| Quantitative variables | 11 | Explain how quantitative variables were handled in the analyses. If applicable, describe which groupings were chosen and why | Methods (‘Study population, data sources, and study design’ & ‘Statistical analysis’) & Table 2 |
| Statistical methods | 12 | (a) Describe all statistical methods, including those used to control for confounding | Methods (‘Statistical analysis’) |
|  |  | (b) Describe any methods used to examine subgroups and interactions | Methods (‘Statistical analysis’), & Table 3 |
|  |  | (c) Explain how missing data were addressed | NA, see Methods (‘Study population, data sources, and study design’) |
|  |  | (d) If applicable, explain how matching of cases and controls was addressed | Methods (‘Study population, data sources, and study design’) |
|  |  | (e) Describe any sensitivity analyses | Methods (‘Statistical analysis’), & Table 3 |
| Results |  |  |  |
| Participants | 13 | (a) Report numbers of individuals at each stage of study—eg numbers potentially eligible, examined for eligibility, confirmed eligible, included in the study, completing follow-up, and analysed<br>(b) Give reasons for non-participation at each stage<br>(c) Consider use of a flow diagram | Figures 3 & 4 |
| Descriptive data | 14 | (a) Give characteristics of study participants (eg demographic, clinical, social) and information on exposures and potential confounders | Table 2 |
|  |  | (b) Indicate number of participants with missing data for each variable of interest | NA, see Methods (‘Study population, data sources, and study design’) |
| Outcome data | 15 | Report numbers in each exposure category, or summary measures of exposure | Results & Table 3 |
| Main results | 16 | (a) Give unadjusted estimates and, if applicable, confounder-adjusted estimates and their precision (eg, 95% confidence interval). Make clear which confounders were adjusted for and why they were included | Results & Table 3 |
|  |  | (b) Report category boundaries when continuous variables were categorized | Mehtods (‘Study population, data sources, and study design')<br>& Table 2 |
|  |  | (c) If relevant, consider translating estimates of relative risk into absolute risk for a meaningful time period | NA |
| Other analyses | 17 | Report other analyses done—eg analyses of subgroups and interactions, and sensitivity analyses | Results & Table 3 |
| **Discussion** |  |  |  |
| Key results | 18 | Summarise key results with reference to study objectives | Discussion, paragraph 1 |
| Limitations | 19 | Discuss limitations of the study, taking into account sources of potential bias or imprecision. Discuss both direction and magnitude of any potential bias | Discussion ('Limitations') |
| Interpretation | 20 | Give a cautious overall interpretation of results considering objectives, limitations, multiplicity of analyses, results from similar studies, and other relevant evidence | Discussion, paragraph 1 |
| Generalisability | 21 | Discuss the generalisability (external validity) of the study results | Discussion ('Limitations') |
| **Other information** |  |  |  |
| Funding | 22 | Give the source of funding and the role of the funders for the present study and, if applicable, for the original study on which the present article is based | Funding |

NA denotes not applicable.

## Results

Figures 3-4 show selection of the study population and Table 2 shows the population’s baseline characteristics. The study population was broadly representative of Qatar’s population.

**Figure 3.**
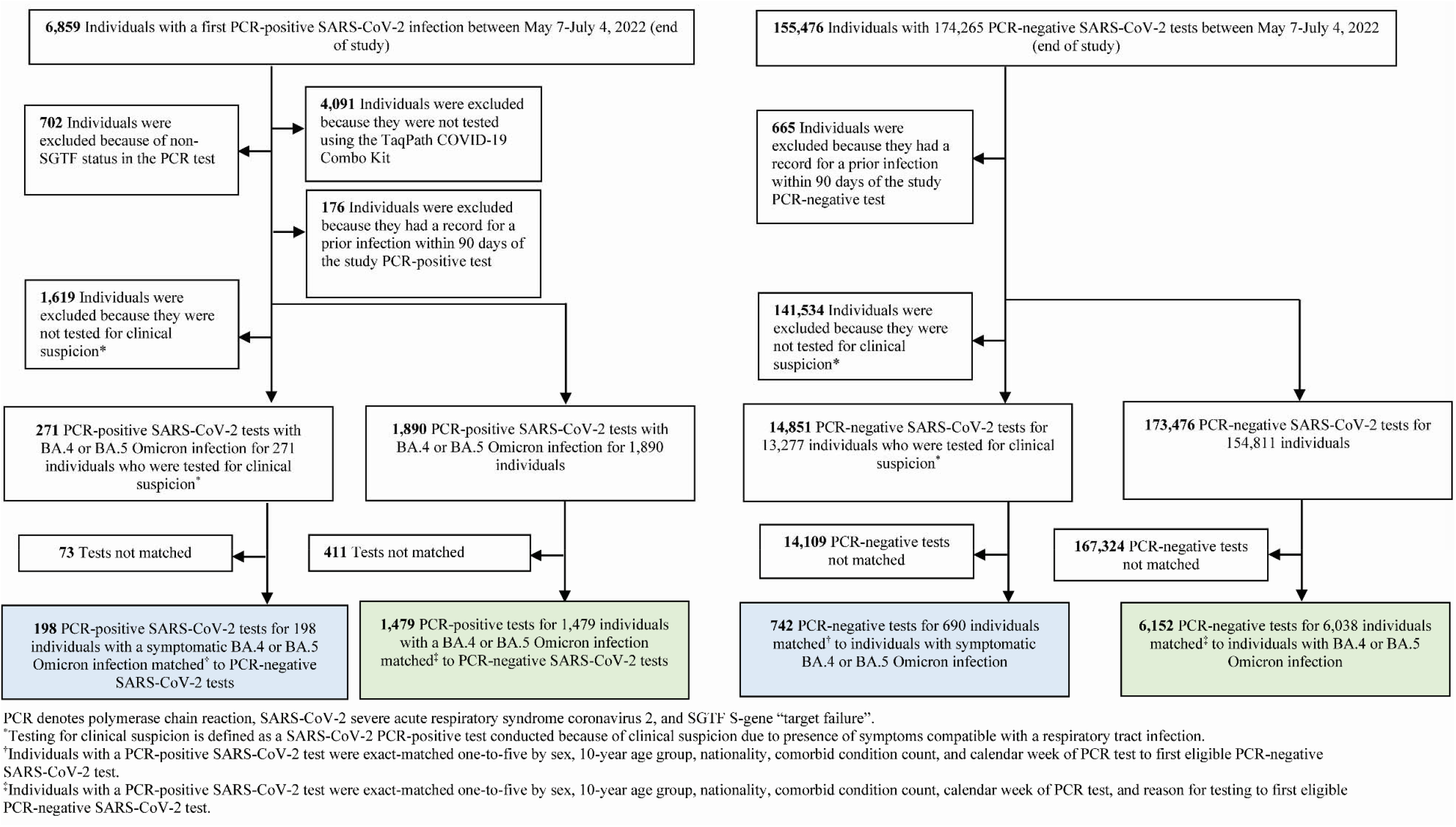
Flowchart describing the population selection process for investigating effectiveness of previous infection in preventing reinfection with the BA.4 or BA.5 subvariants using the S-gene “target failure” diagnosed infections, May 7, 2022-July 4, 2022.

**Figure 4.**
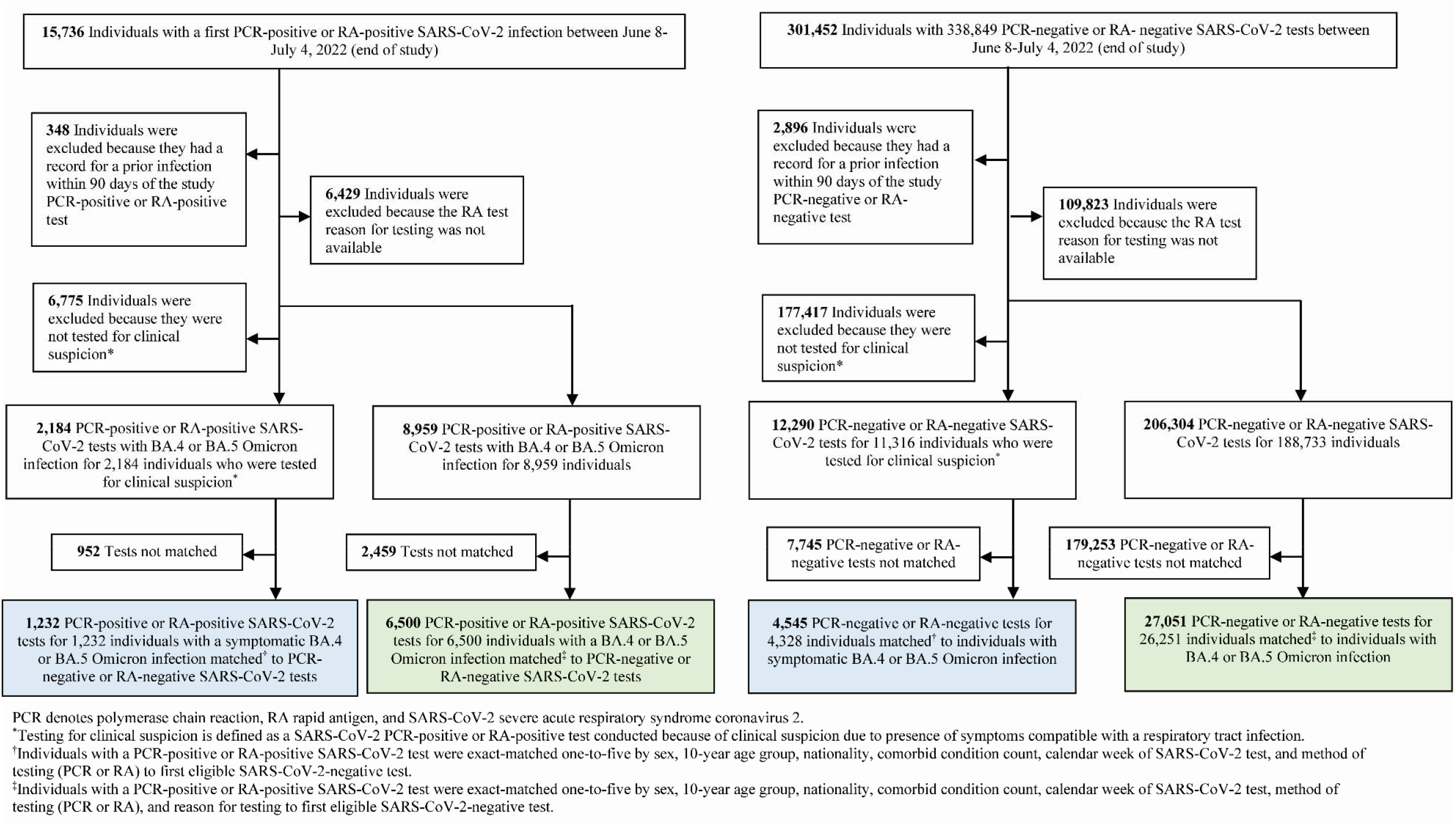
Flowchart describing the population selection process for investigating effectiveness of previous infection in preventing reinfection with the BA.4 or BA.5 subvariants using all SARS-CoV-2 infections diagnosed when BA.4 and BA.5 dominated incidence, June 8, 2022-July 4, 2022.

**Table 2.**
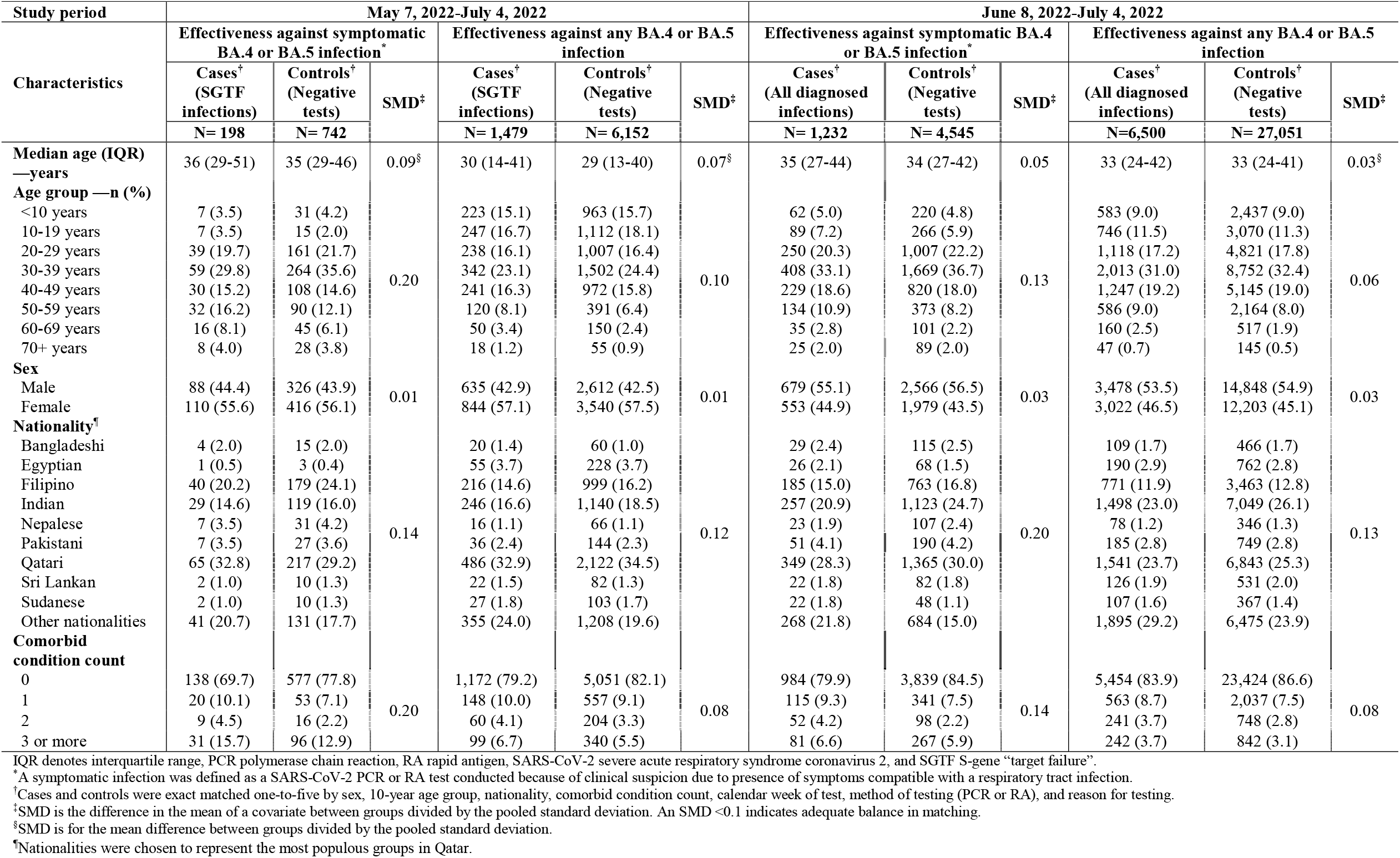
Characteristics of matched cases (SARS-CoV-2-positive tests) and controls (SARS-CoV-2-negative tests) in the analysis assessing effectiveness against symptomatic BA.4 or BA.5 reinfection and in the analysis assessing effectiveness against any BA.4 or BA.5 reinfection regardless of symptoms. The table is generated for the analyses including the S-gene “target failure” infections diagnosed between May 7, 2022 and July 4, 2022, and the analyses including all SARS-CoV-2 infections diagnosed between June 8, 2022 and July 4, 2022, when BA.4 and BA.5 dominated incidence.

Effectiveness of a previous pre-Omicron infection against symptomatic BA.4/BA.5 reinfection was 15.1% (95% CI: -47.1-50.9%), and against any BA.4/BA.5 reinfection irrespective of symptoms was 28.3% (95% CI: 11.4-41.9%) (Table 3). Effectiveness of a previous Omicron infection against symptomatic BA.4/BA.5 reinfection was 76.1% (95% CI: 54.9-87.3%), and against any BA.4/BA.5 reinfection was 79.7% (95% CI: 74.3-83.9%).

**Table 3.** Effectiveness of previous SARS-CoV-2 infection in preventing reinfection with the Omicron BA.4 or BA.5 subvariants using A) S-gene “target failure” infections diagnosed between May 7, 2022 and July 4, 2022, and B) all SARS-CoV-2 infections diagnosed between June 8, 2022 and July 4, 2022, when BA.4 and BA.5 dominated incidence.

| Type of analysis | Cases<sup>†</sup> (SARS-CoV-2-positive tests) |  |  | Controls<sup>†</sup> (SARS-CoV-2-negative tests) |  |  | Effectiveness in % (95% CI)<sup>‡</sup> |
| --- | --- | --- | --- | --- | --- | --- | --- |
|  | Median interval between previous infection and SARS-CoV-2 test (IQR) in days | Previous infection (n) | No previous infection (n) | Median interval between previous infection and SARS-CoV-2 test (IQR) in days | Previous infection (n) | No previous infection (n) |  |
| Primary analyses |  |  |  |  |  |  |  |
| A) Analysis using SGTF status as a proxy for BA.4 or BA.5 |  |  |  |  |  |  |  |
| Effectiveness against symptomatic BA.4 or BA.5 infection* |  |  |  |  |  |  |  |
| Pre-omicron previous infection | 542 (455-713) | 21 | 164 | 498 (427-699) | 77 | 525 | 15.1 (-47.1 to 50.9) |
| Omicron previous infection | 169 (166-175) | 13 | 164 | 167 (159.5-173) | 140 | 525 | 76.1 (54.9 to 87.3) |
| Effectiveness against any BA.4 or BA.5 infection |  |  |  |  |  |  |  |
| Pre-omicron previous infection | 473 (427-628) | 125 | 1,267 | 460 (410-620) | 551 | 4,273 | 28.3 (11.4 to 41.9) |
| Omicron previous infection | 166 (154-173) | 87 | 1,267 | 163 (150-171) | 1,328 | 4,273 | 79.7 (74.3 to 83.9) |
| B) Analysis using any infection during BA.4 and BA.5 dominance |  |  |  |  |  |  |  |
| Effectiveness against symptomatic BA.4 or BA.5 infection* |  |  |  |  |  |  |  |
| Pre-omicron previous infection | 490 (438-685) | 107 | 1,080 | 476 (430-670.5) | 444 | 3,119 | 40.0 (23.9 to 52.7) |
| Omicron previous infection | 167 (161-171) | 45 | 1,080 | 164 (155-171) | 982 | 3,119 | 89.6 (85.5 to 92.6) |
| Effectiveness against any BA.4 or BA.5 infection |  |  |  |  |  |  |  |
| Pre-omicron previous infection | 480 (435-647.5) | 528 | 5,683 | 470 (429-652) | 2,559 | 19,125 | 34.1 (26.9 to 40.5) |
| Omicron previous infection | 167 (159-174) | 289 | 5,683 | 163 (154-171) | 5,367 | 19,125 | 83.8 (81.6 to 85.8) |
| Sensitivity analyses adjusting for vaccination status in conditional logistic regression |  |  |  |  |  |  |  |
| A) Analysis using SGTF as a proxy for BA.4 or BA.5 |  |  |  |  |  |  |  |
| Effectiveness against symptomatic BA.4 or BA.5 infection* |  |  |  |  |  |  |  |
| Pre-omicron previous infection | 542 (455-713) | 21 | 164 | 498 (427-699) | 77 | 525 | 14.9 (-47.5 to 50.9) |
| Omicron previous infection | 169 (166-175) | 13 | 164 | 167 (159.5-173) | 140 | 525 | 76.1 (54.9 to 87.3) |
| Effectiveness against any BA.4 or BA.5 infection |  |  |  |  |  |  |  |
| Pre-omicron previous infection | 473 (427-628) | 125 | 1,267 | 460 (410-620) | 551 | 4,273 | 28.6 (11.8 to 42.2) |
| Omicron previous infection | 166 (154-173) | 87 | 1,267 | 163 (150-171) | 1,328 | 4,273 | 79.7 (74.4 to 84.0) |
| B) Analysis using any infection during BA.4 and BA.5 dominance |  |  |  |  |  |  |  |
| Effectiveness against symptomatic BA.4 or BA.5 infection* |  |  |  |  |  |  |  |
| Pre-omicron previous infection | 490 (438-685) | 107 | 1,080 | 476 (430-670.5) | 444 | 3,119 | 40.3 (24.3 to 52.9) |
| Omicron previous infection | 167 (161-171) | 45 | 1,080 | 164 (155-171) | 982 | 3,119 | 89.6 (85.5 to 92.6) |
| Effectiveness against any BA.4 or BA.5 infection |  |  |  |  |  |  |  |
| Pre-omicron previous infection | 480 (435-647.5) | 528 | 5,683 | 470 (429-652) | 2,559 | 19,125 | 34.9 (27.8 to 41.2) |
| Omicron previous infection | 167 (159-174) | 289 | 5,683 | 163 (154-171) | 5,367 | 19,125 | 84.0 (81.8 to 85.9) |

CI denotes confidence interval, IQR interquartile range, PCR polymerase chain reaction, RA rapid antigen, SARS-CoV-2 severe acute respiratory syndrome coronavirus 2, and SGTF S-gene “target failure”.
<sup>\*</sup>A symptomatic infection was defined as a SARS-CoV-2 PCR or RA test conducted because of clinical suspicion due to presence of symptoms compatible with a respiratory tract infection.
<sup>†</sup>Cases and controls were exact matched one-to-five by sex, 10-year age group, nationality, comorbid condition count, calendar week of test, method of testing (PCR or RA), and reason for testing.
<sup>‡</sup>Effectiveness of previous infection in preventing reinfection was estimated using the test-negative, case-control study design.<sup>2</sup>

Results using all diagnosed infections when BA.4/BA.5 dominated incidence showed higher effectiveness estimates, as expected for an upper bound, but still confirmed the same findings with improved statistical precision (Table 3). Sensitivity analyses adjusting for vaccination status confirmed study results (Table 3).

## Discussion

Protection of a previous infection against BA.4/BA.5 reinfection was modest when the previous infection involved a pre-Omicron variant, but strong when the previous infection involved the Omicron BA.1 or BA.2 subvariant. Importantly, protection of a previous infection against BA.4/BA.5 was lower than that against BA.1/BA.2,^7,8,36^ consistent with BA.4/BA.5’s greater capacity for immune-system evasion than that of BA.1/BA.2.

## Limitations

With the young population of Qatar, our findings may not be generalizable to other countries where elderly citizens constitute a larger proportion of the total population. With the relatively young population of Qatar,^12,37^ the lower severity of Omicron,^38-40^ and the time lag between infection and severe forms of COVID-19, there were too small number of confirmed severe,^41^ critical,^41^ and fatal^42^ COVID-19 cases to estimate *PE*_*S*_ against COVID-19 hospitalization and death due to reinfection. Because also of the relatively small number of infections, it was not possible to generate effectiveness estimates by time since previous infection.

The study is based on SARS-CoV-2 tests done on individuals currently in Qatar. Qatar has a diverse expatriate population, and it is possible that some persons may have had a previous infection diagnosis while traveling abroad to visit family or for vacation, but which would not have been captured in our national databases. However, this is not likely to affect our estimates. It has already been shown that even considerable levels of misclassification of previous infection status had a minimal impact on estimated *PE*_*S*_,^2^ a key strength of the test-negative design.^2^

While matching was done for sex, 10-year age group, nationality, comorbid condition count, calendar week of testing, method of testing (PCR or RA), and reason for testing, this was not possible for other factors such as geography or occupation, as such data were unavailable. However, Qatar is essentially a city state and infection incidence was broadly distributed across neighborhoods. Nearly 90% of Qatar’s population are expatriates from over 150 countries coming here because of employment;^12^ most are craft and manual workers working in development projects.^12^ Nationality, age, and sex provide a powerful proxy for socio-economic status in this country.^12,18-21^ Nationality alone is strongly associated with occupation.^12,18-21^

Matching was done to control for factors known to affect infection exposure in Qatar.^12,18-21^ The matching prescription had already been investigated in previous studies of different epidemiologic designs, and using control groups to test for null effects.^10,13,16,22,23^ These control groups included unvaccinated cohorts versus vaccinated cohorts within two weeks of the first dose,^10,16,22,23^ when vaccine protection is negligible,^43,44^ and mRNA-1273-versus BNT162b2-vaccinated cohorts, also in the first two weeks after the first dose.^13^ These studies have shown that this prescription provides adequate control of the differences in infection exposure.^10,13,16,22,23^ The study was implemented on Qatar’s total population, perhaps thus minimizing the likelihood of bias.

*PE*_*S*_ was assessed using an observational, test-negative, case-control study design,^2^ rather than a cohort study design where individuals are followed up over time. However, the cohort study design applied in earlier analyses to estimate *PE*_*S*_ in the same population of Qatar yielded findings similar to those of the test-negative, case-control study design,^2,6,14,45,46^ supporting the validity of this design in estimating *PE*_*S*_. It even appears that the test-negative study design may be less prone to some forms of bias than the cohort study design.^2^

Nonetheless, one cannot exclude the possibility that in real-world data, bias could arise in unexpected ways, or from unknown sources, such as subtle differences in test-seeking behavior or changes in the pattern of testing. Notwithstanding these limitations, consistent findings were reached in all the different primary and sensitivity analyses.

## Data Availability

The dataset of this study is a property of the Qatar Ministry of Public Health that was provided to the researchers through a restricted-access agreement that prevents sharing the dataset with a third party or publicly. Future access to this dataset can be considered through a direct application for data access to Her Excellency the Minister of Public Health (https://www.moph.gov.qa/english/Pages/default.aspx). Aggregate data are available within the manuscript and its Supplementary information.

## Acknowledgements

We acknowledge the many dedicated individuals at Hamad Medical Corporation, the Ministry of Public Health, the Primary Health Care Corporation, the Qatar Biobank, Sidra Medicine, and Weill Cornell Medicine – Qatar for their diligent efforts and contributions to make this study possible.

The authors are grateful for support from the Biomedical Research Program and the Biostatistics, Epidemiology, and Biomathematics Research Core, both at Weill Cornell Medicine-Qatar, as well as for support provided by the Ministry of Public Health, Hamad Medical Corporation, and Sidra Medicine. The authors are also grateful for the Qatar Genome Programme and Qatar University Biomedical Research Center for institutional support for the reagents needed for the viral genome sequencing. Statements made herein are solely the responsibility of the authors. The funders of the study had no role in study design, data collection, data analysis, data interpretation, or writing of the article.

## Author contributions

HNA co-designed the study, performed the main statistical analyses, and co-wrote the first draft of the article. LJA conceived and co-designed the study, led the statistical analyses, and co-wrote the first draft of the article. HC co-designed the study, performed statistical analyses, and co-wrote the first draft of the article. PVC designed mass PCR testing to allow routine capture of SGTF variants. PT and MRH conducted the multiplex, RT-qPCR variant screening and viral genome sequencing. HY, HAK, and MKS conducted viral genome sequencing. HA contributed to study design. All authors contributed to data collection and acquisition, database development, discussion and interpretation of the results, and to the writing of the manuscript. All authors have read and approved the final manuscript.

